# Minimum material requirements for hand hygiene in community settings: A systematic review

**DOI:** 10.1101/2025.03.12.25323858

**Authors:** Lilly A. O’Brien, Kennedy Files, Jedidiah S. Snyder, Hannah Rogers, Oliver Cumming, Joanna Esteves Mills, Bruce Gordon, Matthew C. Freeman, Bethany A. Caruso, Marlene K. Wolfe

## Abstract

This systematic review assessed the minimum requirements necessary to create an enabling environment for sustained hand hygiene practices: quantity of water and soap, and number, spacing, location, and design of hand hygiene facilities. This review provides evidence for the development of the WHO Guidelines for Hand Hygiene in Community Settings. We searched PubMed, Web of Science, EMBASE, CINAHL, Global Health, Cochrane Library, Global Index Medicus, Scopus, PAIS Index, WHO IRIS, UN Digital Library and World Bank eLibrary, and consulted experts. Eligible studies were observational, in non-healthcare community settings, and reported on at least one of the five categories: (1) quantity of water, (2) quantity of soap, (3) location of hand hygiene materials, (4) number of users or spacing of facilities, (5) considerations for equitable access. Two reviewers independently extracted data from each study and assessed risk of bias using the Mixed Method Appraisal Tool (MMAT). This review identified 37 studies that met inclusion criteria from 28 countries, representing 4 of the 6 WHO regions— (Africa, South-East Asia, the Americas, and Europe). Household settings were the most represented (59% of studies), followed by institutional or school settings (41%) and public establishments (27%). Of the 37 studies, 13 (35%) assessed the relationship between a material requirement and hand hygiene practices. Despite extensive global research on hand hygiene, we found a lack of evidence linking material requirements with handwashing practices in community settings. This review was limited to observational studies, and more data could be derived from experimental studies. Important evidence gaps include the quantity of water and soap needed, the influence of facility location and design on hand hygiene practice, and material needs providing equitable access. Further research is needed to strengthen the evidence base for hand hygiene recommendations and supplement the expert opinion on which many recommendations are currently based.

**What is already known on this topic:** Hand hygiene is an important tool to prevent infectious disease transmission; however, there is little information about what the minimum material requirements are to enable handwashing in community settings to achieve this goal.

**What this study adds:** This systematic review examines the evidence of material needs to enable handwashing in community settings, and the relationship with handwashing-associated outcomes when reported in included studies. It highlights key gaps in the current evidence base, particularly surrounding specifics of quantities and locations of materials. The study proposes that future work collect this data and examine the association between these material needs and hand hygiene practice. This is a necessity for increasing our understanding and providing recommendations on the minimum requirements for effective and sustained hand hygiene in community settings.

**How this study might affect research practice or policy:** This review demonstrates that there is very little evidence providing critical, practical information on what materials are needed to meet handwashing needs in community settings. More detailed and specific reporting of the materials and conditions associated with handwashing, alongside evidence about practice and health and other outcomes, is needed to improve recommendations. Beyond reporting more quantitative details, there was a lack of studies focused on equitable access to handwashing, and further research is needed to specifically address material needs for handwashing for people with disabilities.

## 1. INTRODUCTION

Hand hygiene, whether through handwashing with soap or other methods such as the use of alcohol-based hand rubs (ABHRs), is a critical public health measure. Hand hygiene interventions are relatively inexpensive to implement [1] and can prevent several infectious diseases including enteric and respiratory infections, which account for a large burden of disease [2–4] and high healthcare costs [5]. Establishing global guidelines and recommendations is essential to guide hand hygiene initiatives, protect public health, and strengthen resilient health systems [6].

According to the Ottawa Charter, community settings are where ‘health is created and lived by people within the setting of their everyday life; where they learn, work, play, and love,’ [7] and include domestic, public, and institutional spaces [8]. Guidelines for hand hygiene in healthcare settings are well-established [9–12], and additional guidelines emphasize investing in hand hygiene as a core public health measure [13–16]. Despite the recognized importance of hand hygiene, and the unprecedented prioritization driven by the COVID-19 pandemic [16], gaps and inconsistencies remain in global guidance on specific measures [8]. A recent scoping review identified 51 existing international guidelines and highlighted a lack of consistent evidence-based recommendations and identified four areas where clear recommendations are needed for hand hygiene in community settings: (1) effective hand hygiene; (2) minimum requirements; (3) behavior change; and (4) government measures [8].

Minimum requirements refer to the materials (and in some cases the services and infrastructure) required for effective hand hygiene. Despite the importance of hand hygiene and unprecedented levels of attention and investment during the COVID-19 pandemic, there continues to be insufficient access to basic services for hand hygiene —presence of a handwashing facility with soap and water— especially in many low- and-middle income countries [17–19]. Materials used for hand hygiene often serve additional purposes, such as for cooking, drinking, bathing, and laundry. These competing priorities make allocation of materials to hand hygiene a complex process, and a lack of access to essential materials for all purposes may impede adoption of recommended hand hygiene practices. When examining hand hygiene materials in global recommendations, MacLeod et al. 2023 found that soap and water and ABHR were consistently recommended, and thus likely widely acceptable across community settings [8]. However, recommendations for minimum requirements in key elements such as quantity of soap and water and the number and spacing/location of hand hygiene facilities are inconsistent and lacking across international guidelines, highlighting the need for further research to determine what these requirements are.

The aim of this systematic review was to assess the minimum requirements reported in observational studies of hand hygiene for sustained practice of effective hand hygiene in community settings. The priority question and sub-questions for this review were generated through an extensive consultation process by the WHO with external experts [20,21], following a scoping review of current international guidelines [8].

## 2. METHODS

### 2.1. Research questions

This systematic review sought to assess the following five questions related to minimum requirements for hand hygiene: (a) What quantity of water is required to enable handwashing with soap and water at key moments? (b) What quantity of soap is required to enable handwashing with soap and water at key moments? (c) Where should soap and water or alternatives be located in community settings to enable hand hygiene at key moments? (d) What is the optimal spacing and number of users per hand hygiene facility in household settings and public places to enable hand hygiene with soap and water at key moments? (e) What are the main considerations for ensuring equitable access to minimum material requirements and preventing discrimination in community settings?

### 2.2. Search strategy

This review was pre-registered with PROSPERO (registration number: CRD42023429145) and is reported in accordance with the Preferred Reporting Items for Systematic Reviews and Meta-Analyses (PRISMA) criteria [22](S1 – PRISMA Checklist). This review was part of an integrated protocol for multiple related reviews to synthesize the evidence for effective hand hygiene in community settings [21]. We adopted a two-phased approach for identifying relevant studies. Phase 1 involved a broad search to capture all studies on hand hygiene in community settings that were relevant across multiple related systematic reviews. The outcome of phase 1 was a reduced sample from which further screening, specific to this review, was performed. A full description of the procedures followed for searches, study inclusion, outcomes data collection, analysis, and reporting of the multiple related reviews is presented in the published protocol [21].

This search included studies published between January 1, 1980, and March 29, 2023, and published in English—unless the title and abstract was published in English and/or a non-English language article was referenced in an existing systematic review. We searched 12 peer-reviewed and grey literature databases.

PubMed, Web of Science, EMBASE (Elsevier), CINAHL (EBSCOhost), Global Health (CAB), Cochrane Library, Global Index Medicus, Scopus (Elsevier), Public Affairs Information Service (PAIS) Index (ProQuest) were searched on March 23, 2023 and WHO Institutional Repository for Information Sharing (IRIS), UN Digital Library, and World Bank eLibrary were searched on March 28, 2023 using search terms related to hand hygiene broadly and restrictions on terms related to healthcare settings in the titles. We searched trial registries (International Clinical Trials Registry Platform and clinicaltrials.gov) for trials related to hand hygiene in community settings on March 29, 2023.

In addition, we contacted 35 content experts and organizations, using snowballing methods, from April to May 2023 for information on relevant unpublished literature. Manual searches of reference lists were not completed for this review as we did not identify exiting systematic reviews relevant to the research question.

### 2.3. Selection criteria

For this review, hand hygiene refers to any hand cleansing undertaken for the purpose of removing or deactivating pathogens from hands and efficacious hand hygiene is defined as any practice which effectively removes or deactivates pathogens from hands and thereby has the potential to limit disease transmission [10]. The term community settings included domestic (e.g., households), public (e.g., markets, public transportation hubs, vulnerable populations [e.g., people experiencing homelessness], parks, squares, or other public outdoor spaces, shops, restaurants, and cafes), and institutional (e.g., workplace, schools and universities, places of worship, prisons and places of detention, nursing homes and long-term care facilities) spaces [8]. Studies were excluded if they were in healthcare settings or were animal research. Studies in nursing homes and long-term care facilities were excluded as part of phase 2 screening as these were defined as locations where healthcare is delivered and providing evidence akin to that generated in healthcare settings. There were no geographic restrictions.

Quantitative and were all eligible. Publications not based on empirical research were excluded. Studies were included if the topic of research was on the quantity of water and/or soap required for handwashing with soap at key moments both as recommended and as commonly practiced; the location of soap and water or alternatives to enable hand hygiene at key moments; the spacing and number of users per hand hygiene facility required for handwashing with soap and water at key moments; or considerations (including location and design) leading to harm or inequitable access to handwashing with soap at key moments or discrimination.

We used Covidence software for systematic reviews [23]. In both phases, screening of each article (phase 1 – title and abstract only; phase 2 – title and abstract, then full text review) was performed independently by two reviewers, with discordance between reviewers reconciled by a third reviewer. The stages and related outcomes of the search and screening process are described in the PRISMA flow diagram (S2- PRISMA Flow Diagram).

### 2.4. Data analysis

Two reviewers [LO, KF] independently extracted data using customized data extraction tools (S3 – Covidence extraction and S4 – Outcome extraction) and assessed quality for each article using the Mixed Method Appraisal Tool (MMAT) [24,25]. Any conflicts between reviewers over data extraction and bias assessment were resolved by discussion and checking the source material.

We extracted qualitative data describing the location and design of handwashing facilities, and quantitative data on the amounts of water and soap reported for use and the number of users along with the statistical association between these observations and measures of hand hygiene practice. These findings were summarized narratively and in tables, when sufficient data was reported. We established location categories based on observed data without predetermined classifications due to uncertainty about the locations that would be represented in the data. Due to the scarcity of data, quantitative summaries and meta-analyses were not conducted for this study.

### 2.5. Ethics and Patient/ Public Involvement Statement

Because we reviewed published documents, no ethical approval was required. Patients or the public were not involved directly in the design, or conduct, or reporting, or dissemination plans of our research. This evidence synthesis supports the forthcoming WHO Guidelines for Hand Hygiene in Community Settings; the study questions were developed in broad consultation with a network of key partners. Findings from this review will be disseminated alongside the Guidelines.

### 2.6. Data Availability

All data and extraction templates will be made publicly available upon publication at Figshare (https://figshare.com/projects/Synthesising_the_evidence_for_effective_hand_hygiene_in_community_settings_multiple_related_systematic_reviews/234971).

## 3. RESULTS

### 3.1. Study characteristics of the studies included in this review

We identified 37 unique studies that met inclusion criteria (Table 1). Studies took place in 28 countries, representing 4 of the 6 WHO regions—Africa (n=24 studies), South-East Asia (n=8), the Americas (n=4), and Europe (n=3) (Figure 1 and Table 2). We did not identify any studies from the Eastern Mediterranean region or Western Pacific Region. Domestic, or household, settings were the most represented (in 59% of studies), followed by institutional settings (41%) and public settings (27%). The most common primary outcome was the practice of hand hygiene (62%) followed by different health outcomes (16%). Among institutional settings, primary and secondary schools were most commonly studied, followed by daycare centers and universities. The most frequently described hand hygiene practice was handwashing with soap and water (54%). Table 1 summarizes the characteristics of the studies in greater detail.

**Figure 1.**
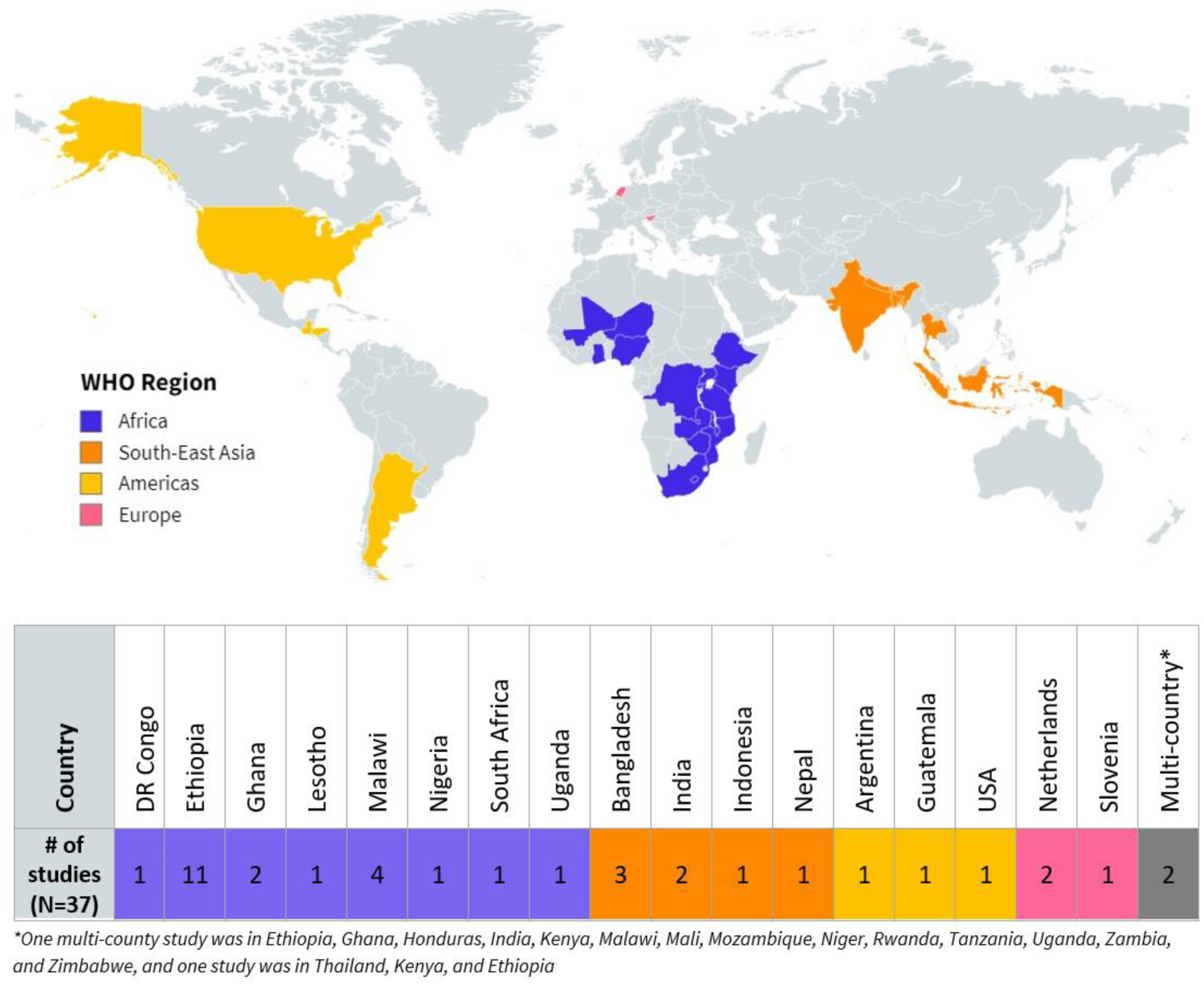
Global distribution of studies reporting on minimum material requirements for hand hygiene in community settings.

**Table 1.** Summary of the included studies.

| **a. Quantity of water available (No identified studies)** |  |  |  |  |  |  |  |
| --- | --- | --- | --- | --- | --- | --- | --- |
| **b. Quantity of soap available** |  |  |  |  |  |  |  |
| **Study ID** | **Community Setting** | **Setting type** | **Country** | **Urban/rural setting** | **Quantity of soap distributed, as presented in the study** | **Handwashing impact assessed?** | **MMAT Score** |
| Biran 2012 | Refugee Camp | Household | Kenya | n/a | No soap distribution | No | 5 |
|  |  |  | Ethiopia | n/a | 1 bar/person/month |  |  |
|  |  |  | Thailand | n/a | 4 bars/person/3 months |  |  |
| Peterson 1998 | Refugee Camp | Household | Malawi | n/a | 240 g/person/month | No | 4 |
| **c. Location of Hand Hygiene facilities** |  |  |  |  |  |  |  |
| **Study ID** | **Community Setting** | **Setting type** | **Country** | **Urban/rural setting** | **Locations assessed, as presented in the study** | **Handwashing impact assessed?** | **MMAT Score** |
| Adane 2018 | Domestic | Households | Ethiopia | Urban | Within or near latrines | No | 5 |
| Admasie 2016 | Domestic | Households | Ethiopia | Urban | In or around the latrine | No | 3 |
| Admasie 2022 | Domestic and Institutional | Households and Schools | Ethiopia | Urban and Rural | At the home (household); In the school compound (school) | Yes | 4 |
| Agaro 2022 | Domestic | Households | Ethiopia | Rural | Near to latrine; Near to household; Not a separate place | No | 5 |
| Aithal 2014 | Domestic | Households | India | Urban and Rural | Toilet; Bathroom; Outside | Yes | 5 |
| Belachew 2018 | Domestic | Households | Ethiopia | Rural | Near to the toilet | No | 4 |
| Berhanu 2022 | Domestic and Institutional | Households and Schools | Ethiopia | Urban and Rural | Next to latrines (household); Presence of a hand washing facility in a student's home (household); In compound within walking distance (school); Within walking distance (school) | No | 4 |
| Besha 2016 | Institutional | Schools | Ethiopia | Urban and Rural | Outside latrine room | No | 4 |
| Chidziwisano 2019 | Domestic | Households | Malawi | Rural | Near the latrine; In the household | No | 5 |
| Fielmua 2021 | Public | Markets and Shops | Ghana | Urban | Handwashing stations around the entrance; Hand sanitizer stations around the entrance | Yes | 5 |
| Green 2007 | Public | Restaurants | USA | Not reported | In worker's sight | Yes | 5 |
| Jubayer 2022 | Domestic | Households | Bangladesh | Not reported | On household premises | No | 5 |
| Kalam 2021 | Domestic | Households | Bangladesh | Urban | Within 10 feet of a latrine or toilet | Yes | 5 |
| Kalumbi 2020 | Domestic | Households | Malawi | Not reported | A fixed handwashing facility (HWF) within the plot/yard; Mobile facility (basin and jug used in multiple locations) | No | 5 |
| Luby 2009 | Domestic | Households | Bangladesh | Rural | Inside or near the toilet facility; Inside or near the kitchen; Elsewhere in the yard; No specific place; Outside the yard | Yes | 3 |
| Melaku 2023 | Institutional | Schools | Ethiopia | Not reported | Toilets; Lounges and dining areas; School yard | No | 5 |
| Moffa 2021 | Domestic | Households | See footnotes for countries<sup>1</sup> | Rural | Within 5 meters of the food preparation area; Within 5 meters of the latrine; Neither location | No | 5 |
| Mugambe 2021 | Public | Supermarkets | Uganda | Urban | Entrance of supermarket | No | 5 |
| Mwapasa 2022 | Institutional | Childhood development centers | Malawi | Peri-urban | At the food preparation area; Near the latrines | Yes | 5 |

| Natnael 2022 | Public | Barbershops and beauty salons | Ethiopia | Not reported | Presence of hand-washing facility with water and soap;<br>Presence of hand-washing facility that is convenient and user friendly in/near the barbershop/the beauty salon | Yes | 4 |
| --- | --- | --- | --- | --- | --- | --- | --- |
| Prevolsek 2021 | Public | Food vendor stalls | Slovenia | Urban | Suitably located washbasins | No | 5 |
| Rao 2021 | Domestic | Households | Guatemala | Not reported | Within 10 meters of sanitation facility | No | 5 |
| Ray 2011 | Domestic and Institutional | Households and Schools | India | Urban | Near the toilets (household); Near the toilet (school) | No | 4 |
| Serra 2015 | Institutional | Daycare centers | Argentina | Urban | In the day care center | No | 4 |
| Shrestha 2021 | Domestic | Households | Nepal | Not reported | Inside toilet; Inside kitchen; Within courtyard; Other | No | 4 |
| Sibiya 2013 | Institutional | Schools | South Africa | Urban and Rural | Inside their toilets; At the center of schools, about 100 meters from the toilet | No | 3 |
| Sondari 2020 | Institutional | Schools | Indonesia | Not reported | Facilities present | Yes | 5 |
| Tessema 2021 | Domestic | Households | Ethiopia | Not reported | Hand washing facility available | Yes | 5 |
| van Beeck 2016 | Institutional | Daycare centers | Netherlands | Urban and Rural | All facilities were present in playroom (soap, sink, and towel) | No | 5 |
| Wami 2022 | Institutional | Schools | Nigeria | Urban | At the entrance of the school | No | 5 |
| Zomer 2013 | Institutional | Daycare centers | Netherlands | Urban and Rural | At least one sink in each classroom | No | 4 |
| **d. Number of facilities per area or user** |  |  |  |  |  |  |  |
| Study ID | Community Setting | Setting type | Country | Urban/rural setting | Number of facilities per area or user, as presented in the study | Handwashing impact assessed? | MMAT Score |
| George 2022 | Public and Institutional | Fifteen indoor public location types | Democratic Republic of the Congo | Urban | Multiple hand sinks | Yes | 4 |
| Green 2007 | Public | Restaurants | USA | Not reported | Multiple handwashing stations | No | 5 |
| van Beeck 2016 | Institutional | Daycare centers | Netherlands | Urban and Rural | 2 sinks with taps per playroom | No | 5 |
| Zomer 2013 | Institutional | Daycare centers | Netherlands | Urban and Rural | 2 sinks, towel facilities, and soap facilities per classroom per 2 caregivers | Yes | 4 |
| **e. Design considerations** |  |  |  |  |  |  |  |
| Study ID | Community Setting | Setting type | Country | Urban/rural setting | Design of facility, as presented in the study | Handwashing impact assessed? | MMAT Score |
| Kalam 2021 | Domestic | Households | Bangladesh | Urban | Presence of a permanent handwashing facility in the home | Yes | 5 |
| Kebede 2022 | Domestic | Households | Ethiopia | Urban and Rural | Observed, fixed place; Observed, mobile place | Yes | 3 |
| Letuka 2021 | Public | Markets | Lesotho | Urban | Buckets to pour water over hands | No | 5 |
| Sondari 2020 | Institutional | Schools | Indonesia | Not reported | Hand washing facilities with permanent buildings | No | 3 |
| Steiner-Asiedu 2011 | Domestic and Institutional | Households and Schools | Ghana | Urban | Sinks with running water (households and schools); Receptors for communal handwashing (households and schools); Plastic containers with a tap (households and schools) | No | 4 |
| Wami 2022 | Institutional | Schools | Nigeria | Urban | Standpipe | No | 5 |
| Zomer 2013 | Institutional | Daycare centers | Netherlands | Urban and Rural | Paper towels or fabric towels; Soap pump or bar soap or soap dispenser | Yes | 4 |

<sup>1</sup> Ethiopia, Ghana, Honduras, India, Kenya, Malawi, Mali, Mozambique, Niger, Rwanda, Tanzania, Uganda, Zambia, and Zimbabwe

**Table 2.** Characteristics of the included studies.

| **Descriptive characteristics of studies\*** | **Total**<br>**n (%)** | **Quantitative**<br>**n (%)** | **Mixed methods**<br>**n (%)** |
| --- | --- | --- | --- |
| **Total number of studies** | **37 (100%)** | **24 (65%)** | **13 (35%)** |
| **Region** |  |  |  |
| African Region | 24 (65) | 13 (54) | 11 (85) |
| South-East Asian Region | 8 (22) | 7 (29) | 1 (8) |
| Region of the Americas | 4 (11) | 3 (13) | 1 (8) |
| European Region | 3 (8) | 3 (13) | - |
| Eastern Mediterranean Region | - | - | - |
| Western Pacific Region | - | - | - |
| Multiple regions | 2 (5) | 1 (4) | 1 (8) |
| **Urban/Rural** |  |  |  |
| Urban | 12 (32) | 6 (25) | 6 (46) |
| Rural | 5 (14) | 4 (17) | 1 (8) |
| Peri-urban | 1 (3) | - | 1 (8) |
| Other: Refugee camp | 2 (5) | 1 (4) | 1 (8) |
| Both urban and rural | 8 (22) | 7 (29) | 1 (8) |
| Unspecified | 9 (24) | 6 (25) | 3 (23) |
| **Setting** |  |  |  |
| Domestic (Households) | 22 (59) | 16 (67) | 6 (46) |
| Institutional | 15 (41) | 10 (42) | 5 (38) |
| Daycare center | 4 (11) | 3 (13) | 1 (8) |
| Primary and Secondary Schools | 10 (27) | 6 (25) | 4 (31) |
| Universities | 1 (3) | 1 (4) | - |
| Public | 10 (27) | 5 (21) | 5 (38) |
| Restaurant | 2 (5) | 1 (4) | 1 (8) |
| Food vendor stalls | 1 (3) | - | 1 (8) |
| Barbershops and beauty salons | 2 (5) | - | 2 (15) |
| Markets | 4 (11) | 3 (13) | 1 (8) |
| Places of worship | 1 (3) | 1 (4) | - |
| **Sex of primary research population** |  |  |  |
| Female | 3 (8) | 2 (8) | 1 (8) |
| Male | - | - | - |
| Unspecified or both male and female | 34 (92) | 22 (92) | 12 (92) |
| **Primary outcome studied** |  |  |  |
| Hand hygiene | 23 (62) | 14 (58) | 9 (69) |
| Health outcome | 6 (16) | 6 (25) | - |
| Diarrheal diseases | 4 (11) | 4 (17) | - |
| Nutrition | 1 (3) | 1 (4) | - |
| Neglected tropical diseases | 1 (3) | 1 (4) | - |
| WASH access or KAP | 5 (14) | 1 (4) | 4 (31) |
| COVID-19 KAP | 3 (8) | 3 (13) | - |
| Food hygiene | 2 (5) | 1 (4) | 1 (8) |
| **Hand hygiene practice** |  |  |  |
| Handwashing with soap and water | 20 (54) | 10 (42) | 10 (77) |
| Handwashing with water | 4 (11) | 1 (4) | 3 (23) |
| Handwashing with soap and water or water only | 4 (11) | 3 (13) | 1 (8) |
| Hand washing with soap and water or ash | 2 (5) | 2 (8) | - |
| Handwashing with soap and water or ABHR | 2 (5) | 2 (8) | - |
| Handwashing with ABHR | 1 (3) | - | 1 (8) |
| More than 2 methods | 5 (14) | 4 (17) | 1 (8) |
| Handwashing - unspecified | 2 (5) | 2 (8) | - |
| **Study quality rating (mean)** | **4.46** | **4.38** | **4.62** |

*\*Descriptive statistics for region, setting, primary outcome studied, and hand hygiene practice represent variables that were multi-select, so the sum of sections may be over the study total.*

### 3.2. Quality of the studies included in this review

The mean study quality was 4.46 overall indicating good quality, 4.38 for quantitative descriptive studies (n=24), and 4.62 for mixed methods studies (n=13). All studies received a score of 3 or higher. S5 – MMAT Assessment presents quality appraisal scores for each study included.

### 3.3. a. Quantity of water

We did not identify any observational studies that reported on the quantity of water available or required to enable hand hygiene at key moments in community settings.

#### 3.3.b. Quantity of soap

No studies reported the quantity of soap available or required to enable handwashing in community settings, but two studies (Table 1) reported the quantity of soap distributed in refugee settings [26,27]. We excluded *a priori* studies that noted only presence or absence of soap, as our research questions sought to determine quantity rather than presence. Peterson 1998 reported that 240 g of soap were distributed per person per month at a refugee camp in Malawi and Biran 2012 reported that no soap distribution occurred at a camp in Kenya, one bar of soap per person was distributed at a camp in Ethiopia, and four bars of soap per person was distributed every three months at a camp in Thailand. Biran (2012) also reported on the prevalence of both handwashing with water only and handwashing with soap by refugee camp, but neither study carried out an analysis of the association between the quantity of soap distributed and hand hygiene behavior. The observed percentage of potential handwashing events in Biran (2012) that occurred with both water and soap was low overall, but was highest in Ethiopia (19%), followed by Kenya (17%), and then Thailand (15%).

#### 3.3.c. Location of hand hygiene facilities

Thirty-one studies reported on the observed or self-reported location of hand hygiene facilities in domestic settings (households), institutional settings (schools and daycare centers), or public settings (markets, supermarkets, restaurants, food vendors, and barber and beauty shops) (Table 1) [28–57]. Nine individual studies reported the associations between location of hand hygiene facilities and hand hygiene practices (Table 3 and S6 - Facility location associations).

**Table 3.**
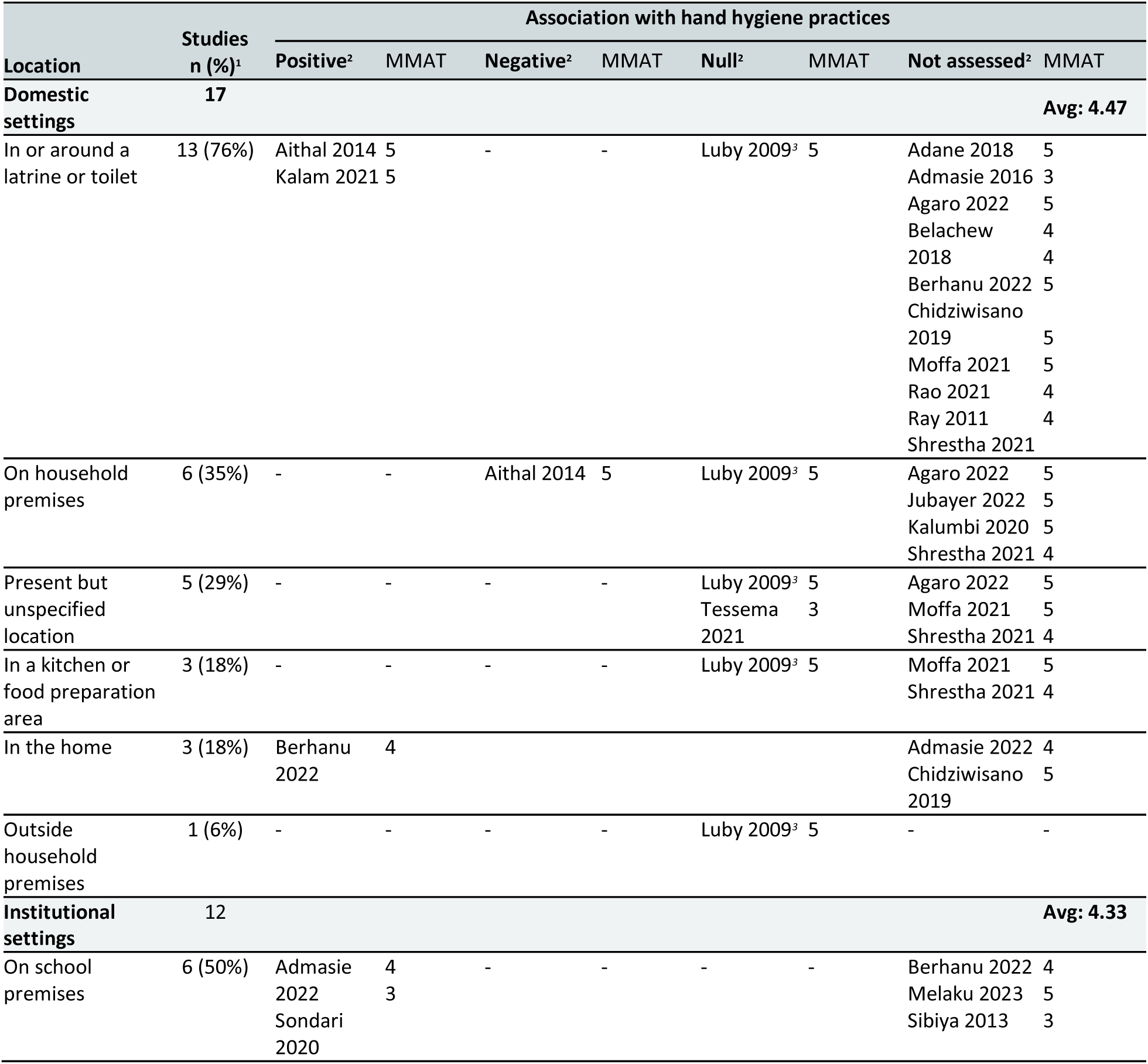
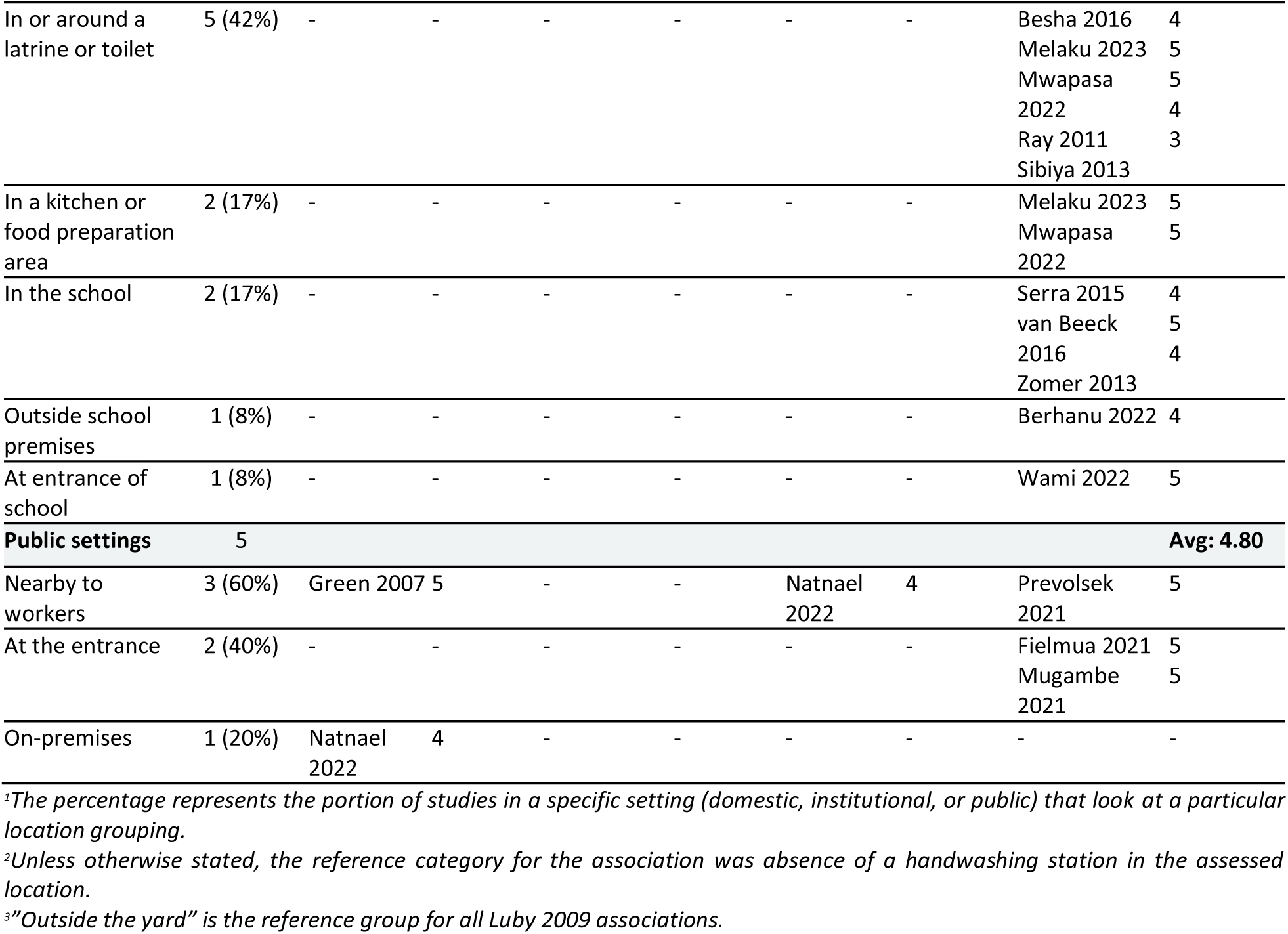
Association of locations of hand hygiene facilities with hand hygiene practices.

Hand hygiene facility locations were categorized into the following categories for domestic settings (categories were based on extracted data): in or around a latrine or toilet; in a kitchen or food preparation area; in the home (not specified to be in/around a latrine/toilet or in a kitchen area); on household premises (either in the yard or near the home); outside household premises, but available to participants; unspecified location, but available to participants. Locations of hand hygiene facilities in institutional settings were categorized as: in or around a latrine or toilet; in a kitchen or food preparation area; in a school (not specified to be in/around a latrine/toilet or in a kitchen area); on school premises (either noted to be in the school yard or present without a specified location); outside school premises, but available to students; or at the entrance of the school. Resulting locations reported from public settings were categorized as: nearby to workers (in-sight or easily accessible); at the entrance of a public establishment; or on-premises of a public establishment.

Of the studies in domestic settings, 76% recorded whether a handwashing station was observed “in or around a latrine or toilet” (Table 3). The next most common location assessed was the presence or absence of a hand hygiene facility “on household premises” (35% and 29% of studies, respectively). Two studies found positive associations between the location of “in or around a latrine or toilet” and hand hygiene practices compared to no facility at the location. Aithal 2014 found that a higher proportion of mothers practiced hand hygiene when the wash area was in or near the toilet compared to outside the toilet [58]. Similarly, Kalam 2021 found a higher proportion practiced hand hygiene when there was a handwashing facility within 10 feet of a toilet compared to the absence of a toilet in that location [36]. Berhanu 2022 also found positive associations between reported hand hygiene practices and having a facility in a student’s home [32]. Luby 2009 reported null associations between hand hygiene practices for the location “inside or near the toilet” and other locations, specifically when “outside the yard” was the reference group [38]. Tessema 2021 reported null associations between “hygiene practices” and the presence or absence of a facility, noting that their measure of “hygiene practices” was an aggregate measure that also included non-hand hygiene measures (S6 - Facility location associations) [51].

In the institutional settings, 50% of studies considered the simple presence or absence of a facility, and 42% considered a location of in or around a latrine or toilet (Table 3). Two studies found a positive association between having hand washing facilities on school premises and improved hand hygiene practices when compared to having no facilities available; improved hand hygiene was defined based on two different composite scoring systems that both considered performance of handwashing at critical times, using appropriate materials, and frequency (S6 - Facility location associations) [30,50].

In public settings, one study (Green 2007, food vendors) found a positive association between having a facility nearby to a worker and “appropriate” hand washing practices (as defined by the paper, including removal of gloves, handwashing with soap and water, and drying) being used [57], but another (Natnael 2022, barber shops and beauty salons) reported a null association for the same location [43]. Conversely, Natnael 2022 found a positive association between increased hand hygiene practices and the presence of a hand hygiene facility compared with no facility present (S6 - Facility location associations).

#### 3.3.d. Number of facilities per area or users

Four studies reported on factors related to the number of facilities per area or per users, and two studies reported on associations of these factors with hand hygiene practices (Table 1) [52,54,57,59]. Green 2007 reported that food workers were more likely to “appropriately” wash their hands (definition above) when multiple hand sinks were available in a restaurant compared to 1 or fewer sinks [57]. Zomer 2013 reported that an increased number of towel facilities per caregiver was positively associated with hand hygiene compliance, but the number of sink facilities and soap facilities per caregiver were not associated with hygiene outcomes (S7 - Number of facilities associations) [54].

#### 3.3.e. Design considerations of hand hygiene facilities

Seven studies reported on the design of the hand hygiene facilities specifically, and three studies reported on associations of these factors with hand hygiene practices (Table 1) [36,50,53,54,60–62]. Both Kalam 2021 and Kebede 2022 found that the odds of users practicing hand hygiene were higher when the facility was a fixed or permanent design compared to a mobile facility [36,60]. Zomer 2013 reported that caregivers in a daycare center were more likely to wash their hands when there were only paper towels available compared to either fabric towels or both paper towels and fabric towels [54]. They did not find an association between types of soap facilities available (soap pump, soap bar, or soap dispenser) and handwashing behaviors (S8 - Design associations).

## 4. DISCUSSION

Our review of the minimum requirements necessary to enable sustained, effective handwashing in community settings found a lack of evidence to describe the material requirements in terms of quantity of soap and water, location, spacing, and design of handwashing stations - and even less evidence relating these material needs to hand hygiene practices. Although the literature was too sparse for consensus to emerge from the available data, one area in which our findings were consistent was that the presence of any hand hygiene facility in community settings supports sustained handwashing. Overall, the lack of sufficient evidence across the literature regarding these minimum requirements and their association with hand hygiene is a critical gap, as this information is important in informing future recommendations and interventions to improve hand hygiene.

### **a.** Quantity of Water

We found no data that specified the quantity of water available and used for handwashing in the studies included in this review. While during the screening process we found many studies reporting on the presence or absence of water available for handwashing, these did not provide any information on water volume. Presence/absence information may provide insight into overall access to water however it does not address scarcity nor enable quantification of the amount of water needed to enable effective handwashing as defined by WHO guidelines. Future studies should quantify the amount of water available and used for handwashing, even if roughly estimated, to help determine water supply needs for handwashing. This is especially important for planning water supplies in public and institutional settings that serve large populations. Although recommendations for water quantity requirements exist based on expert opinion and estimations, recommendations on quantity are few and discordant [8]. Further data is needed to align and support evidence-backed recommendations.

We aimed to determine how the observed quantities of water at a handwashing station are associated with the frequency or quality of the handwashing practice, but we did not find studies reporting this association. Quantity may influence the quality of handwashing practice. For example, an individual may have access to enough water to wash their hands, but if there is not excess water, they may choose to refrain from washing their hands and conserve the water for another time or purpose [63–66]. Studies have shown that hand hygiene interventions are only effective at reducing mortality in non-water scarce settings and when there is a predictable water supply available. This epidemiological evidence supports the critical role of water in hand hygiene and emphasizes the importance of understanding water supply and scarcity beyond presence and absence [67,68]. This is especially pertinent for domestic settings where many competing priorities for water may exist. In a companion review by our team focused on barriers and enablers to hand hygiene, we identified that “water prioritization” for other activities and “water supply quantity” were both noted as barriers to handwashing, specifically in domestic settings [69]. A recent WHO publication also reported insufficient empirical evidence to define minimum quantities of water necessary for cooking and hygiene, including personal and food hygiene [70]. Through expert opinion, it was determined that 20 L/person/day would allow for hand and face washing in addition to drinking, cooking, and food hygiene, and the SPHERE Handbook providing standards for humanitarian response indicates that people must be provided with 15L/person/day to meet basic needs including hygiene [71]. It also noted that the amount of water used for domestic purposes was influenced by accessibility, continuity, reliability, and price of water, with water use being prioritized for drinking and cooking purposes. Given these complexities and the lack of empirical data to attach water quantities required for each of these needs, further research is critical to establish minimum water requirements that support hand hygiene both independently and within the context of broader domestic needs.

### **b.** Quantity of soap

We found a lack of observational data on the quantities of soap available to users in community settings. The presence or absence of soap was reported in many studies, though this alone was not enough to determine the quantity of soap present or needed for sustained handwashing. Two observational studies reported on quantity of soap distributed in refugee camp settings, though they did not explore the relationship between soap quantity present in households and hand hygiene practices [27,72]. Although not intervention studies, these studies also reflect a unique context in which hygiene materials are regularly distributed to individuals in humanitarian settings. Interestingly, one study assessed the reported priority use of the distributed soap and determined that laundry was the highest priority, followed by bathing, then either dishes or handwashing in all three countries (Thailand, Kenya, and Ethiopia) [27]. This same order of prioritization for soap was identified in Uganda [73]. As with water, the domestic setting presents multiple competing needs for soap that must be taken into account when determining recommendations for the quantity needed. Further research is needed to determine how much soap is necessary for effective and sustained handwashing in diverse settings, particularly in low-resource environments. This question of how much soap is needed to enable sustained handwashing in community settings may be more clearly examined in intervention studies with prescribed soap distribution or in an observational study primarily dedicated to this research question.

### **c.** Location of hand hygiene facility

Evidence from the literature indicates that the presence of a designated facility in any location supports the practice of hand hygiene, however the data was insufficient to generate consensus or provide more detail on where facilities should be located. The included studies were primarily conducted in domestic and school settings rather than public settings. Hand hygiene facilities were most frequently assessed “in or near latrine/toilet facilities” (76% of studies in domestic settings and 42% of studies in institutional settings). Most studies that examined the association between location of handwashing facilities and hand hygiene practices focused on the presence or absence of a facility in key locations, often broadly defined. Of these, the majority found a positive association between hand hygiene practices and the presence of a hand hygiene facility in any location, when compared to not having a facility at all. This finding is supported by a meta-analysis from national surveys that found hand washing with soap was about two times more likely to occur when there was a designated facility present compared with no designated facility present [74] The aforementioned companion review by our team also identified “hand hygiene facilities location” as a barrier/enabler to hand hygiene across domestic, institutional, and public settings - noting that inconvenient location of handwashing facilities limited hand hygiene practice, and the inverse enabled practice [69]. It is well established that hand hygiene practices decrease as distance between a handwashing facility and the home or latrine increases [75]. This suggests that, while simply having a designated facility available promotes handwashing, there are certain locations that are more desirable than others for handwashing facilities. These locations are likely those in proximity to key hand hygiene moments (e.g., before meals, after using the toilet, or entering a public space) in specific locations (e.g., near food preparation areas, bathrooms, or entrances). However, given the number of potentially different locations that require hand hygiene in key moments in a singular domestic, institutional, or public setting, further studies are needed to determine which specific locations of hand hygiene facilities have more significant effects on hand hygiene practices, particularly in low-resource settings where the number of facilities built and supplied may be limited.

### **d.** Number of hand hygiene facilities per area or user

When it comes to the number of hand hygiene facilities required for sustained handwashing, it is unclear from the available data whether having multiple facilities has a significant effect on hand hygiene practices or how many may be required per user. We found few studies reporting on the number of hand hygiene facilities per area or user; of a total of four studies, only two reported associations with hand hygiene practicesand each were in different settings and with mixed results. One study reported that the presence of multiple sinks in a restaurant area was associated with workers practicing “appropriate” handwashing (defined above) [57], while another study in a daycare center setting found that multiple sinks per caregiver had no association with handwashing [54]. Though these measures are not identical, they can be interpreted as measures of accessibility of hand hygiene facilities to workers. No conclusions can be drawn from this small sample of data. The Zomer et al. 2013 study in the daycare center, though finding null associations between number of sinks per caregiver and soap facilities per caregiver, found a significant association between the number of towel facilities per caregiver and handwashing practices. This study provides a good model of considering the different components of hand hygiene materials separately for their individual effects on hand hygiene that future studies should employ.

### **e.** Design of hand hygiene facility

We sought to understand primary considerations for ensuring equitable access to minimum requirements to hand hygiene, yet we found no literature that looked at material availability or its relationships with hand hygiene through an equity lens. Further research on equity is warranted given that a lack of access is known to disproportionately affect certain groups, including women and girls and people with disabilities [18,76]. We must note that thorough determination of considerations for equitable access to minimum requirements will vary greatly throughout different environmental, socio-cultural, and economic contexts. We did identify two studies that considered the design of hand hygiene facilities that may impact accessibility and usability. These found that having a permanent or fixed hand hygiene facility was beneficial for hand hygiene practices compared to a mobile facility, suggesting that the specifics of hand hygiene facility design are important to consider. Therefore, recommendations on how to assess individual contexts to determine what would ensure equitable access to these requirements may be most useful and should be published to improve this body of literature.

### Strengths and Limitations

This review was part of an integrated protocol for multiple related reviews, which included an exhaustive search strategy encompassing multiple databases and grey literature sources, and a two-phased approach to identify relevant literature of hand hygiene in community settings. To our knowledge, this is the first review to systematically search the literature for observational studies addressing the minimum requirements for effective hand hygiene in community settings. Owing to the comprehensive nature of our search, this review includes studies from diverse global contexts from four of six WHO regions and across all three community setting types (domestic, institutional, and public). Despite these strengths, several limitations should be acknowledged. Our scope was focused on observational data to investigate these relationships in natural community settings, but additional data on the minimum requirement required to sustain handwashing could be derived from experimental studies. Most significantly, because there is insufficient data in the literature regarding material requirements for hand hygiene specific to our research questions, we were not able to establish a consensus across different contexts and hand hygiene outcomes for our research questions. Further, we were unable to perform planned meta-analyses or draw firm conclusions due to the lack of data. This limitation may have been eased by broadening our scope to include experimental data, though the intention of gathering data on natural community settings would then have been compromised. Despite—and due to—the lack of data, the review has highlighted key gaps in existing research that are key to determining the needs for minimum requirements for hand hygiene.

## 5. CONCLUSION

This systematic review highlights key gaps in our understanding of the minimum requirements necessary for effective and sustained hand hygiene in community settings. While many recommendations exist on specific quantities of water and soap required, as well as the ideal locations, number of users, and design considerations for hand hygiene facilities, these recommendations are generally based on expert opinion and estimates rather than robust evidence. There is a clear need for evidence to guide these recommendations in the future to enhance the progress of the hand hygiene sector and improve hand hygiene practices globally. Field research can be costly and challenging, and so opportunities to collect and report this kind of data within funded projects should be considered. Future research focusing on hand hygiene practice should seek to integrate observations about materials requirements for handwashing, quantifying and describing these materials and (where possible) relating them to the hand hygiene outcomes of interest in these studies.

## Authors’ contributions

OC, JEM, and BG conceived the review and designed the specific research questions. BAC and MW are the guarantors of the review. LAO and MKW conceived and designed the specific analysis strategy described here. LAO led design of the data extraction process with MKW and JSS support. HR led the literature search, and KF supported the screening process. LAO and KF conducted the data extraction. LAO led the analysis, led manuscript writing, and oversaw revisions. JSS drafted portions of the manuscript. All authors read and approved the final version of the manuscript.

## Acknowledgements

The authors would like to extend their gratitude to all individuals and institutions whose contributions made this systematic review possible. Special thanks to the team of screeners who contributed substantially to the review process as part of phase 1 and phase 2 screening (Erika Canda, Lanelle Nwogalanya, Jordan Honeycutt, Michael Horner-Ibler, Rosemary Madaki, Kennedy Files, Nick An, Erin LaFon, Shahreen Hussain, Norah McKinley, Josef Zhao, Kainalu Bailey). We are also grateful to the authors of the studies included in this review for their important contributions to the field.

## Funding statement

This work was supported by the World Health Organization.

## Competing interests

None declared.

## Patient Involvement Statement

Patients or the public were not involved directly in the design, or conduct, or reporting, or dissemination plans of our research. This evidence synthesis supports the forthcoming WHO Guidelines for Hand Hygiene in Community Settings; the study questions were developed in broad consultation with a network of key partners. Findings from this review will be disseminated alongside the Guidelines.

## Notes

**Funding:** This work was supported by the World Health Organization (PO number: 203046633).

### Competing Interest Statement

The authors have declared no competing interest.

### Funding Statement

This work was supported by the World Health Organization (PO number: 203046633).

